# *Hippocrates-o1*: A Guideline-Aware, Orchestrated, Self-Refining Protocol for Specialty-Specific Clinical Reasoning

**DOI:** 10.64898/2025.12.04.25341678

**Authors:** Benlu Wang, Elizabeth Schaefer, Nicole Aguirre, Juncheng Huang, Xiawei Li, Santhi Raj Kolamuri, Lee Ying, Xinyue Li, Grace Chao, Sheryl S. Ang, Prashanth Vallabhajosyula, Harlan Krumholz, Karen E. Gibbs, Sara I. Pai, Eric B. Schneider, Arman Cohan, Chin Siang Ong

**Author notes:** **Corresponding Author:** Chin Siang Ong, MBBS, PhD, MPH, Assistant Professor of Surgery, Division of Surgical Outcomes, Surgery Center for Health Services and Outcomes Research, Department of Surgery, Yale School of Medicine.

## Abstract

**Background:** Clinical decision support requires language models that provide guideline-aligned, context-aware reasoning with clear justification. Many existing benchmarks emphasize multiple-choice or short-form question answering and mainly capture factual recall rather than longitudinal clinical reasoning from extended clinical notes. Hippocrates-o1 is a family of domain-tailored clinical reasoning pipelines that combine structured prompts, guideline-informed retrieval, and iterative self-refinement across oncology, general surgery, and vascular surgery.

**Methods:** Real-world head and neck cancer cases were drawn from the MIMIC-IV-Note database, with a subset (n=20) randomly selected for detailed annotation. Six physicians adjudicated treatment phase and intent using structured criteria and rated model outputs. For each case, we generated outputs using both a general-purpose baseline model (*VanillaLLM*) and our oncology-specific reasoning model, *Hippocrates-Karkinos-o1*. Experts evaluated the outputs across five dimensions on a scale of 1 to 5: Clinical Knowledge Application, Contextual Understanding, Reasoning Transparency, Chain-of-Thought Quality, and Hallucination Audit. Overall Reasoning was the mean of domain scores. To explore whether the approach could extend beyond oncology, we also processed inguinal hernia and aortic aneurysm cases through *Hippocrates-Chirurgos-o1* and *Hippocrates-Angios-o1* domain adaptations.

**Results:** Across paired ratings, *Hippocrates-Karkinos-o1* improved Overall Reasoning from 3.40±0.90 to 4.00±0.73 (p<0.001). Domain scores increased for Clinical Knowledge Application (2.87±1.20 to 3.70±1.03), Contextual Understanding (3.48±0.95 to 3.98±0.95), Hallucination Audit (3.90±1.32 to 4.74±0.76), Reasoning Transparency (3.45±1.02 to 3.86±0.87), and Chain-of-Thought Quality (3.32±1.04 to 3.69±1.00), all p≤0.001. Surgical and vascular adaptations showed parallel qualitative improvements.

**Conclusions:** The Hippocrates-o1 protocol improved reasoning fidelity, guideline alignment, and factual grounding relative to a general-purpose model and generalized across oncology, surgery, and vascular care. Orchestrated retrieval and self-refinement provide a reproducible template for evaluating and enhancing clinical reasoning in medical AI.

## Introduction

Large language models (LLMs) are increasingly used in clinical informatics and medical decision support, offering new capabilities for summarization, reasoning, and knowledge synthesis. As these systems approach clinical application, there is growing recognition that model performance must be assessed not only by accuracy, but also by the transparency and contextual soundness of the reasoning process, as highlighted in a recent commentary on advanced LLMs, such as GPT-5^1^. Reliable evaluation of reasoning quality is essential for safe and interpretable deployment in patient care.

Most current evaluations of medical LLMs rely on question-answering (QA) benchmarks^2,3^, such as MedQA, PubMedQA, or MultiMedQA, and training approaches designed to optimize performance on these benchmarks^4^. These datasets often only measure factual recall and shallow reasoning in narrowly defined tasks. Although useful for benchmarking general knowledge, such tests do not reflect the complexity of real clinical reasoning, where decisions depend on longitudinal context, evolving treatment phases, and patient-specific factors^5,6^. High performance on knowledge-recall QA datasets does not necessarily translate into clinically reliable or guideline-consistent decision support^7^.

In real-world settings, clinicians interpret unstructured documentation, integrate multiple diagnostic and therapeutic variables, and justify choices according to established standards of care^8^. AI systems intended for clinical deployment must therefore demonstrate not only correctness of recommendations but transparency of rationale and adherence to evidence-based guidelines^9–11^. These requirements highlight the limitations of generic models and underscore the need for tailored, domain-specific reasoning systems that reflect the structure and decision logic of particular medical specialties^12^.

To address this gap, we developed *Hippocrates*-*o1*, a reproducible, guideline-aware reasoning protocol designed to evaluate and support tailored reasoning in medical AI. The protocol combines guideline-informed retrieval with iterative self-refinement, producing explicit, multi-step rationales that reflect both factual accuracy and contextual understanding. The term “o1” denotes the first released specification of this orchestrated reasoning pipeline. Unlike single-task QA evaluations, *Hippocrates-o1* applies structured case-based assessments drawn from real clinical notes, enabling consistent evaluation across diverse specialties and model architectures.

In this study, we present *Hippocrates-o1*, a reproducible orchestration and evaluation protocol for evaluating clinical reasoning across multiple medical domains. Using representative use cases from oncology, general surgery, and vascular surgery, we applied the protocol to compare two base model architectures (Llama and OSS-20B) under two reasoning protocols (*VanillaLLM* versus *Hippocrates-o1*). The findings demonstrate that *Hippocrates-o1* provides a guideline-aware, interpretable, and reproducible basis for assessing reasoning quality, supporting the development of tailored and clinically aligned AI systems.

## Methods

### Data Source and Case Selection

We conducted a retrospective evaluation of clinical reasoning quality using de-identified patient discharge summaries from the MIMIC-IV-Note v2.2 database^13^, linked via subject to structured diagnosis and procedure codes from the MIMIC-IV v3.1 database^14^. The primary analysis focused on head and neck cancer cases, with exploratory analyses in general surgery and vascular surgery. For the oncology domain (*Hippocrates-Karkinos-o1*), we identified discharge summaries for patients with head and neck cancer using ICD codes^15^ (Supplementary Appendix, Table S1). From this cohort, we randomly selected 20 representative cases that contained sufficient detail about oncologic treatment history, intent, and follow-up in order to support productive annotation. For the general surgery domain (*Hippocrates-Chirurgos-o1*), we constructed a cohort of adult patients undergoing inguinal hernia repair using ICD codes (Supplementary Appendix, Table S2). For the vascular domain (*Hippocrates-Angios-o1*), we identified patients undergoing vascular surgery using ICD codes (Supplementary Appendix, Table S3). For each of these domains, we selected 20 representative cases on a basis similar to the oncology domain for reasoning comparison.

### Overview of the *Hippocrates-o1* Architecture

The *Hippocrates-o1* protocol (Figure 1) is a structured, three-stage clinical reasoning pipeline designed to transform unstructured clinical documentation into guideline-aligned recommendations supported by transparent reasoning. The architecture emphasizes reproducibility and clinical interpretability by enforcing clear boundaries between stages and by using explicit input-output specifications. Across all stages, the goal is to convert noisy free-text notes into stable, verifiable decisions and concise surgical or management summaries applicable to multiple clinical domains, including inguinal hernia repair, head and neck cancer, and aortic aneurysm management.

**Figure 1.**
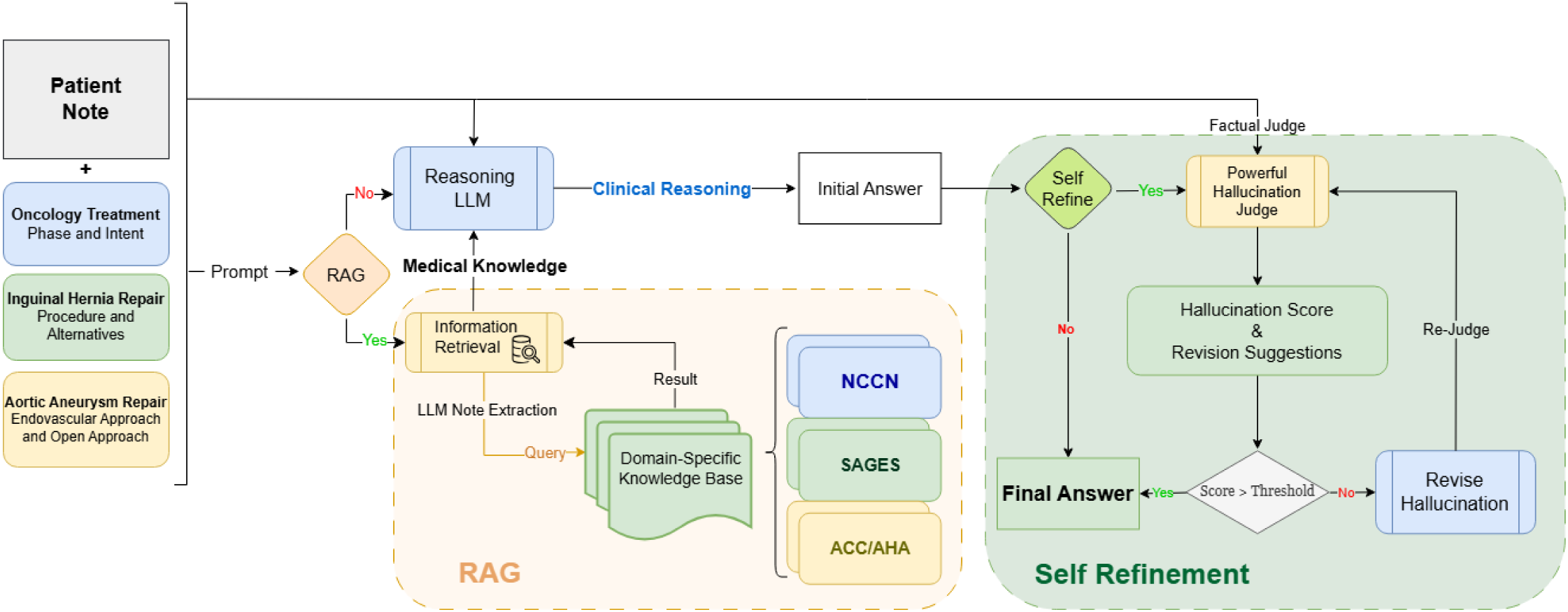
Architecture of the *Hippocrates-o1* Reasoning Protocol. The *Hippocrates-o1* protocol employs a unified reasoning architecture consisting of three stages: (1) initial interpretation, (2) guideline-aware evidence retrieval, and (3) structured self-refinement and consolidation. Each domain adaptation (*Karkinos*, *Chirurgos*, and *Angios*) operates within this shared structure. A patient note is combined with a prompt, for example requesting a procedure summary, possible alternatives, and supporting clinical reasoning. The Reasoning LLM produces an initial answer using domain knowledge. If additional information is required, a retrieval-augmented generation (RAG) component queries domain-specific knowledge bases such as oncology, surgical, or vascular corpora. The output is then refined through an iterative evaluation process in which a hallucination judge assigns a factuality score and provides revision guidance until a final, guideline-aligned output is obtained.

### Stage 1: Initial interpretation and structured case representation

In the initial interpretation stage, the model processes each selected raw clinical note with a domain-tailored prompt to generate a provisional structured output. This output follows a fixed schema to ensure completeness (Supplementary Appendix, B.1/2/3). When the model provides explicit reasoning cues, these are extracted and recorded as part of an interpretable and auditable reasoning trace to enhance transparency and clinical safety. For example, DeepSeek-R1 is one of such models that encloses its intermediate chain-of-thought in a <rawcot>…</rawcot> block. We constructed the pipeline to preserve this block verbatim in a dedicated reasoning-trace field that is outputted alongside the final answer, so that the raw CoT can always be easily located and inspected separately from, but in parallel with, the final structured output.

### Stage 2: Guideline-based evidence retrieval

After this initial interpretation stage, the evidence-retrieval stage augments the structured case representation with authoritative clinical guidance using a retrieval-augmented generation (RAG) approach. This is an important step in ensuring the reliability of output by grounding the model outputs on clinical guidelines. In particular, standard guideline documents^16–18^ are segmented into small, coherent passages and indexed for rapid retrieval. Each passage retains provenance information, including the originating guideline and its section or chapter. A retrieval query is constructed by combining the clinical note with a task-specific clinical question (Table 1, *Representative Task*). When needed, a brief automated extraction step filters the note to identify clinically relevant elements, such as aortic diameters or tumor staging details, thereby improving the relevance of retrieved guidance (Supplementary Appendix, B.5). The system selects the most pertinent guideline excerpts and incorporates them into the model’s context in a concise format, ensuring that downstream reasoning remains grounded in established clinical standards.

**Table 1.**
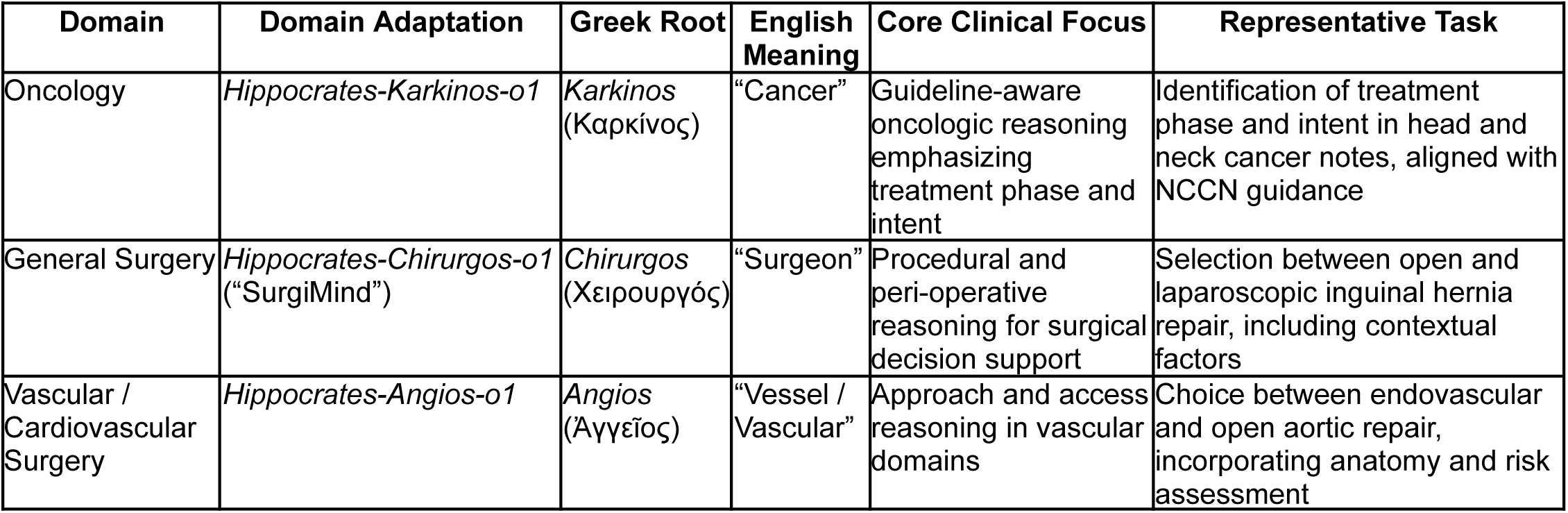
Domain Adaptations within the *Hippocrates-o1* Protocol. The *Hippocrates-o1* protocol extends a shared reasoning protocol across multiple clinical domains. Each domain adaptation integrates domain-relevant knowledge and task-specific objectives while maintaining the same reasoning, retrieval, and refinement structure. The table summarizes the Greek roots, English meanings, clinical focus, and representative reasoning tasks for each domain adaptation: *Karkinos* (oncology), *Chirurgos* (“SurgiMind,” general surgery), and *Angios* (vascular and cardiovascular surgery).

### Stage 3: Structured self-refinement and hallucination auditing

Following retrieval, a structured self-refinement stage is performed to minimize hallucinations. An external auditing component, implemented using a separate Gemini-2.5-pro model, evaluates each part of the provisional output against the source note and assigns a 1 to 5 hallucination support score (higher = better alignment with the note) together with a brief, evidence-grounded explanation (Supplementary Appendix, B.4), accompanied by a brief explanation identifying unsupported or contradictory elements when present. Any segment that does not achieve the maximum hallucination-support score (5), indicating some degree of unsupported content, is routed into targeted revision steps. During revision, the model receives the original note, the prior output, and the auditor’s comments, and is instructed to apply only minimal edits needed to resolve the flagged issues while preserving supported content. The refinement loop is capped at three iterations with early stopping, terminating once all answers meet the hallucination threshold or an iteration yields no further changes. All audit results, revisions, and decision changes are logged to ensure traceability. The final output is a stable structured rationale that summarizes the clinical intent, key decision elements, and supporting guideline-anchored evidence, integrating the reasoning traces produced earlier in the pipeline into a transparent, clinically accountable decision record.

### Model backbones and deployment

The *Hippocrates-o1* architecture is model-agnostic and can operate with different underlying model backbones. In this study, two open-weight models were integrated into the orchestrated pipeline, a Llama-3.1-8B-Instruct model^19^ and a GPT-OSS-20B^20^ model. These models served as interchangeable engines for inference across experiments, with specific pairings varying by domain adaptation so that evaluations reflected the reasoning protocol rather than any single model family. Selecting open-weight models enables local or institution-controlled execution of the reasoning pipeline, allowing protected clinical text to remain within secure environments. This allows the workflow to operate without external cloud-hosted models and supports privacy-preserving workflows aligned with institutional data governance requirements.

### Domain Adaptations within the *Hippocrates-o1* Protocol

The *Hippocrates-o1* protocol extends a shared orchestrated reasoning protocol across multiple clinical domains. Each domain adaptation integrates domain-specific guidelines and task objectives while maintaining the same retrieval and refinement structure. Table 1 summarizes the Greek roots, meanings, and representative tasks for the three adaptations. The *Karkinos* domain adaptation focuses on guideline-aware oncologic reasoning for head and neck cancer, with emphasis on identifying treatment phase and intent in alignment with clinical guidelines. The *Chirurgos* domain adaptation (“SurgiMind”) targets general surgery decision support, with the primary use case involving adult inguinal hernia repair and the contextual factors that influence the selection of open or laparoscopic approaches. The *Angios* domain adaptation focuses on cardiovascular surgery, supporting decisions between endovascular and open aortic repair by incorporating anatomic and peri-operative risk considerations. All adaptations follow a common schema for inputs, intermediate representations, and outputs, enabling direct comparison of reasoning quality across domains.

### Quantitative Assessment Using Structured Evaluation Rubric

The primary quantitative evaluation focused on the *Karkinos* domain adaptation. Twenty de-identified head and neck cancer discharge summaries were randomly sampled from the MIMIC-IV-Note database^13^ and converted into standardized prompts requesting extraction and interpretation of oncologic treatment details, including treatment phase and intent. For each case, paired outputs were generated using the same model backbone under two reasoning configurations: *VanillaLLM*, in which the model received only the clinical prompt and produced a single direct answer, and *Hippocrates-o1*, in which the model was embedded within the orchestrated reasoning pipeline with guideline-informed retrieval and self-refinement before producing a structured rationale.

Six physicians (NA, JH, XL, SRK, GC, SSA), including attending-level clinicians, served as expert raters. All six raters independently evaluated all 20 paired case outputs under both reasoning configurations. Each rater scored outputs according to the rubric in Table 2, which operationalizes five domains of reasoning quality: Clinical Knowledge Application, Contextual Understanding, Reasoning Transparency, Chain-of-Thought Quality, and Hallucination Audit. Anchors for the 1 to 5 scale are defined within the rubric to standardize interpretation across raters. An Overall Reasoning score was then calculated as the mean of the five domain scores. Scores were entered through Label Studio^21^, which enforced completion of all rubric fields. For each domain, the mean of independent rater scores served as the final quantitative value.

**Table 2.** Standardized Clinical Reasoning Evaluation Rubric.

| Dimension | Definition | Scoring Criteria (1–5) |
| --- | --- | --- |
| 1. Reasoning Transparency | Assesses whether the medical rationale is explicitly stated, logically structured, and traceable. | 1 = No justification or contradictory<br>2 = Minimal or fragmented justification<br>3 = Basic logical sequence with at least one explicit connector<br>4 = Mostly structured and interpretable with minor omissions<br>5 = Fully explicit, clear, and stepwise reasoning. |
| 2. Chain-of-Thought Quality | Evaluates clarity, completeness, and multi-step inferential progression. | 1 = No coherent chain or contradictory<br>2 = Single-step reasoning only<br>3 = Partial chain with two or more connected steps<br>4 = Mostly complete, coherent, and clinically meaningful<br>5 = Comprehensive, multi-step reasoning including rationale and alternatives. |
| 3. Clinical Knowledge Application | Measures alignment with correct medical knowledge, standards, and guideline-supported decision making. | 1 = Unsafe or contraindicated recommendations<br>2 = Multiple significant knowledge errors<br>3 = Mostly correct but missing key criteria<br>4 = Correct and guideline-aligned with minor refinements missing<br>5 = Fully correct, guideline-based, and clinically optimal. |
| 4. Contextual Understanding | Evaluates incorporation of patient-specific data, comorbidities, risks, and situational factors. | 1 = Entirely generic<br>2 = Minimal tailoring<br>3 = Incorporates at least one relevant contextual factor<br>4 = Mostly personalized with only minor omissions<br>5 = Fully tailored and integrates multiple relevant clinical and contextual elements. |
| 5. Hallucination Audit | Scores factual grounding strictly against the patient note. Higher score indicates fewer unsupported claims. | 1 = Critical hallucinations with safety risk<br>2 = Major unsupported claims<br>3 = Multiple unsupported elements with moderate impact<br>4 = Minor unsupported detail with low impact<br>5 = No hallucination and fully supported or inferable. |

### Qualitative Assessment

To examine generalizability beyond oncology, we conducted an exploratory qualitative assessment of the *Chirurgos* and *Angios* domain adaptations. Representative cases were selected using the same preprocessing pipeline and domain-adapted prompting templates used for the *Karkinos* domain. Paired *VanillaLLM* and *Hippocrates-o1* outputs were generated using the same underlying model backbone assigned to that domain. Qualitative review (BW, NA, CSO) focused on clarity of rationale structure, integration of guideline content, attention to patient-specific features, and identification of unsupported statements. Observed patterns were summarized in Table 3 and in the specialty-specific supplemental tables (Supplementary Appendix, Tables S4 to S6).

**Table 3.** Representative Case Comparisons for *Karkinos-o1*, *Chirurgos-o1*, and *Angios-o1*: *VanillaLLM* versus *Hippocrates-o1* Reasoning.

| Case | *VanillaLLM* | *Hippocrates-o1* |
| --- | --- | --- |
| Post-chemoradiation tonsillar SCC mucositis | 1. Misclassifies the patient as still in an active chemoradiation phase rather than post-treatment surveillance.<br>2. Conflates acute post-treatment toxicity with ongoing therapy and underuses later context (mucositis, PEG feeds, neutropenia, discharge course).<br>3. Implies ongoing radiation despite documentation that chemo-radiation is complete, with only partially transparent reasoning. | 1. Correctly frames the case as post-treatment surveillance after definitive cisplatin-based chemoradiation.<br>2. Uses the full chart trajectory (mucositis, febrile neutropenia, PEG feeding, recovery and discharge) to anchor the phase of care.<br>3. Avoids speculative further-therapy claims and offers a clearer stepwise narrative from evidence to conclusion. |
| Chronic right groin pain after prior inguinal mesh repair | 1. Identifies open right groin exploration with mesh removal and Bassini repair but underemphasizes ilioinguinal neurectomy and neuropathic pain.<br>2. Proposes laparoscopic or tissue repair alternatives without clearly linking them to prior mesh, neurectomy needs or multimorbidity.<br>3. Overweights DVT-related bleeding risk and speculates that alternatives are less ideal without clear chart support, with limited use of transplant status, immunosuppression, chronic opioids and psychiatric comorbidities. | 1. Recognizes open mesh removal and Bassini repair and more carefully discusses laparoscopic TAPP as a minimally invasive alternative and potential pain benefit.<br>2. Explicitly incorporates transplant status, immunosuppression, multiple prior surgeries and pain/psychiatric comorbidities into perioperative risk.<br>3. States when the chart does not specify the surgical rationale and refrains from inventing one, yielding calibrated, chart-grounded reasoning. |
| Ascending aortic aneurysm (hemiarch replacement) | 1. Correctly identifies ascending aorta and hemiarch replacement with a 30 mm Dacron graft under DHCA as open repair and mentions TEVAR/hybrid options generically.<br>2. Uses aneurysm size and involvement to justify open surgery without explicitly invoking guideline thresholds or key risk modifiers.<br>3. Underuses prior carotid-dissection stroke and family sudden cardiac death, giving correct but high-level, less anatomy-specific and less transparent reasoning. | 1. Frames the same operation as the prototypical open approach for proximal ascending/arch aneurysm.<br>2. Describes the endovascular alternative as an ascending/arch stent-graft with concrete limitations (landing zones, endoleak, branch-vessel compromise).<br>3. Explicitly cites ACC/AHA thresholds and explains earlier intervention based on risk modifiers and arch/root involvement, integrating prior cerebrovascular event, family sudden death and valve/root changes into a stepwise argument. |

### Statistical Analysis

The primary endpoint was the difference in mean Overall Reasoning score between the *Karkinos* domain adaptation and the corresponding *VanillaLLM* outputs. For each case, the average rating across raters for the *Hippocrates-o1* output and the corresponding *VanillaLLM* output was calculated, and their paired difference was used as the analytic unit. Paired two-sided t tests were applied to these differences for the Overall Reasoning score and for each rubric domain. A significance threshold of p < 0.05 was used. Values are reported as mean ± standard deviation. Figure 2 presents domain-level mean scores for each reasoning configuration, standard error of the mean, and the associated p values.

**Figure 2.**
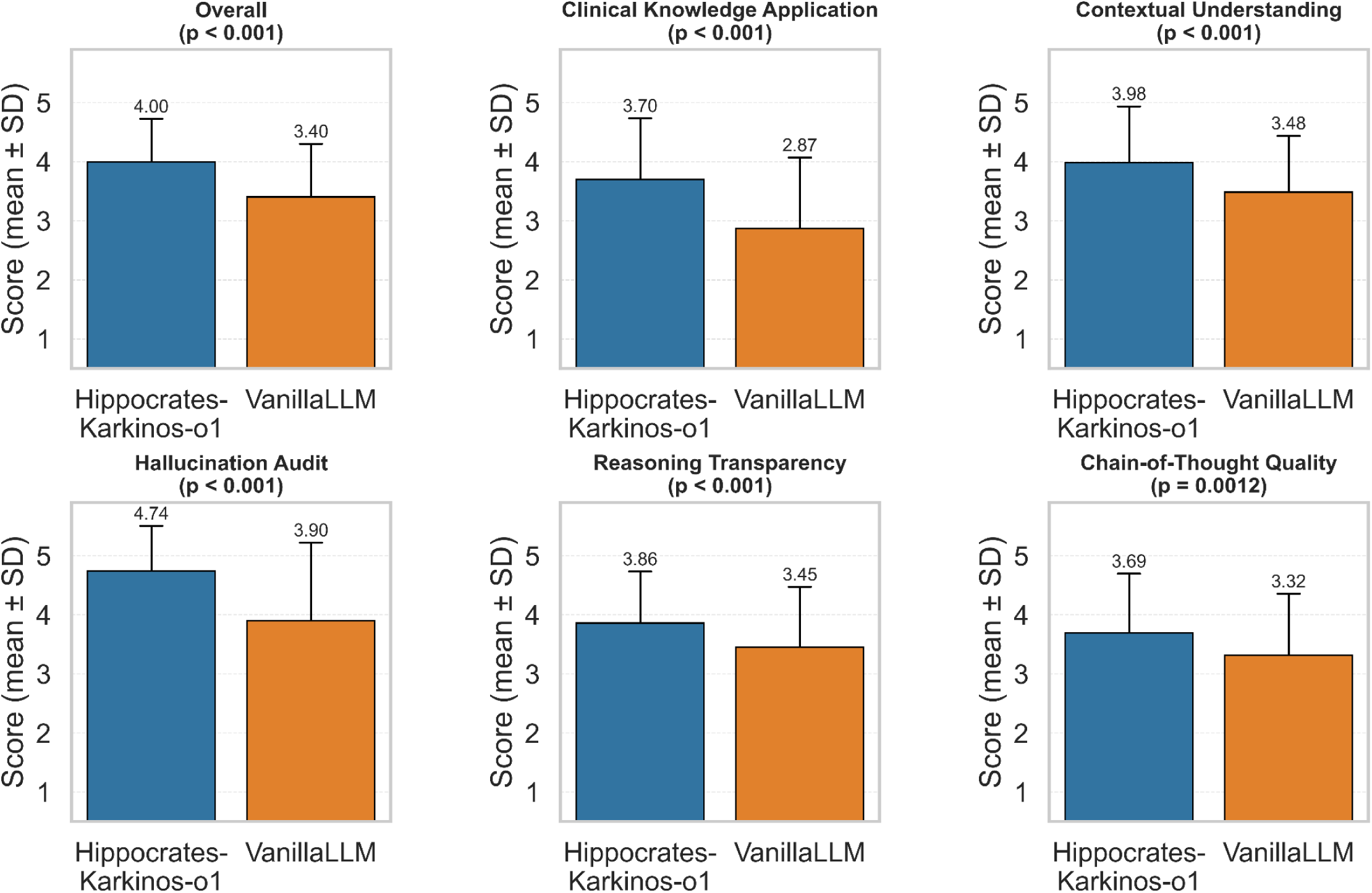
Performance comparison of *Hippocrates-Karkinos-o1* domain adaptation versus a baseline LLM (“*VanillaLLM*”) across expert-rated clinical reasoning domains. The *Hippocrates-Karkinos-o1* oncology adaptation demonstrates significantly higher mean scores with lower standard deviation across all evaluated domains: Overall score, clinical knowledge application, contextual understanding, hallucination audit, reasoning transparency, and chain-of-thought quality. All comparisons reach statistical significance with reported p values below conventional thresholds, indicating improved domain aligned reasoning fidelity, reduced reasoning errors, and greater transparency relative to *VanillaLLM*.

## Results

### Quantitative Results (*Hippocrates-Karkinos-o1* Domain Adaptation)

Across paired case evaluations, *Hippocrates-Karkinos-o1* demonstrated significant quantitative improvements in structured reasoning quality, compared with the baseline *VanillaLLM*. Mean overall reasoning scores increased from 3.40 ± 0.90 to 4.00 ± 0.73 (p < 0.001), indicating both higher reasoning fidelity and reduced score variability among annotators. Gains were observed across all five predefined evaluation domains: *Clinical Knowledge Application* (2.87 ± 1.20 to 3.70 ± 1.03, p < 0.001), *Contextual Understanding* (3.48 ± 0.95 to 3.98 ± 0.95, p < 0.001), *Hallucination Audit* (3.90 ± 1.32 to 4.74 ± 0.76, p < 0.001), *Reasoning Transparency* (3.45 ± 1.02 to 3.86 ± 0.87, p < 0.001), and *Chain-of-Thought Quality* (3.32 ± 1.04 to 3.69 ± 1.00, p = 0.001).

### Cross-Domain and Cross-Model Generalization

Across the *Chirurgos* and *Angios* domain adaptations (Table 1), qualitative and quantitative patterns paralleled those observed in *Karkinos* domain adaptation. Both *Chirurgos* and *Angios* demonstrated clearer rationale structure, reduced hallucination frequency, and improved contextual justification relative to their respective baseline models. These results indicate that the structured retrieval-and-refinement protocol improved reasoning quality independent of disease type or procedural domain. Comparable gains were also observed across alternative model architectures, including *OSS-20B*, confirming that the observed enhancements are attributable to the *Hippocrates-o1* reasoning protocol itself rather than any single underlying model.

Together, these findings demonstrate that the *Hippocrates-o1* architecture generalizes effectively across clinical specialties, supporting reproducible, guideline-aware reasoning from surgical to oncologic and vascular decision contexts.

### Qualitative Reasoning Comparison

Qualitative analysis further highlighted the nature of these improvements. Qualitative review of three representative cases (Table 3; Supplemental Tables S4-S6), drawn from the 20-case qualitative sets in each domain, illustrated how these improvements manifested in practice. *VanillaLLM* frequently produced unsubstantiated conclusions, such as asserting “the treatment was curative” without clear justification, reflecting hallucinated or over-confident reasoning. In contrast, *Hippocrates-Karkinos-o1* appropriately acknowledged uncertainty when evidence was incomplete, producing statements like “the treatment intent is unclear based on available documentation”. This restraint improved both factual reliability and interpretability.

Nevertheless, reviewer feedback noted that the model’s reasoning remained partially generic, often omitting relevant patient-specific details that could enrich contextual understanding and Chain-of-Thought depth. Some limitations were traced to the underlying discharge summaries, which frequently lacked comprehensive oncologic information. These findings suggest that model reasoning quality may be further improved through enhanced data context and structured documentation inputs.

## Discussion

This study demonstrates reproducible improvement in clinical reasoning quality, transparency, and guideline alignment using the *Hippocrates-o1* domain adaptations. Compared with a general-purpose LLM, *Hippocrates-o1* consistently produced clearer, evidence-based rationales with fewer unsupported assertions. Quantitatively, this was reflected in higher domain scores across all five reasoning categories, while qualitative assessment showed improved calibration, particularly reduced hallucination of treatment intent and a greater tendency to acknowledge uncertainty when documentation was incomplete.

The central innovation of this work lies in establishing *Hippocrates-o1* as a cross-domain, reproducible reasoning protocol for medical AI. By combining guideline-informed retrieval with iterative self-refinement, the protocol moves beyond factual recall toward structured reasoning evaluation. Existing QA-style benchmarks such as MedQA or MultiMedQA quantify factual recall but fail to capture the longitudinal, context-dependent reasoning fundamental to clinical practice^3,22^. Our findings suggest that reasoning-based evaluation better reflects how clinicians synthesize data, weigh evidence, and justify decisions, and may ultimately guide both regulatory assessment and safe model deployment.

Rather than supplanting expertise, *Hippocrates-Karkinos-o1* enhances it. Oncologic decision-making requires integration of evolving guidelines, heterogeneous data, and nuanced patient context, cognitive tasks that challenge even experienced clinicians. A system that retrieves relevant evidence, reasons through treatment intent, and exposes its logic can reduce cognitive load while preserving human accountability. In this way, the *Hippocrates-o1* protocol embodies collaborative intelligence: physicians and AI co-reasoning to deliver safer, more consistent, and more transparent care.

This augmentation is particularly valuable in community and resource-limited settings, where oncologists may lack ready access to subspecialty expertise or the latest NCCN updates. By embedding guideline-awareness and transparent reasoning into every inference, the *Hippocrates-o1* family of models could democratize access to evidence-based oncology, reducing variability between academic and non-academic centers.

Beyond oncology, generalizability of the protocol to multiple domains confirms the scalability of this approach. *Hippocrates-Chirurgos-o1*, applied to inguinal hernia repair decisions (open vs laparoscopic, mesh vs non-mesh), and *Hippocrates-Angios-o1*, focused on aortic aneurysm management (endovascular vs open repair), demonstrated consistent improvements in rationale clarity, contextual accuracy, and reduction of unsupported conclusions. These results underscore that the *Hippocrates-o1* protocol generalizes across both disease and procedural reasoning, forming a unified foundation for transparent AI across medical specialties. Quantitative scoring combined with qualitative feedback provides a multidimensional view of how models reason, critical for aligning AI behavior with clinical, ethical, and regulatory standards.

Limitations include the small annotated sample size and reliance on discharge summaries that incompletely represent longitudinal care. Human evaluation, though essential for construct validity, introduces subjectivity that future automated rubric scoring may mitigate.

Future directions include extending *Hippocrates-o1* reasoning to additional cancer types and specialties, integrating multi-modal data, and advancing to *Hippocrates-o2* with enhanced retrieval precision and self-explanation, and incorporating auditable intermediate reasoning traces. Embedding such reasoning protocols within electronic health systems could fundamentally reshape oncology, creating an ecosystem where human and artificial reasoning operate synergistically to deliver guideline-concordant, patient-centered care at scale.

## Conclusion

*Hippocrates-o1* is a structured clinical reasoning protocol that integrates guideline-informed retrieval and iterative self-refinement, representing our first released specification of an orchestrated, guideline-aware reasoning pipeline. It establishes a reproducible, multi-domain orchestration and evaluation protocol for clinical AI, providing a standardized and interpretable benchmark that supports tailored assessment across multiple medical domains.

## Data Availability

All data analyzed in this study are available through controlled-access repositories and from the authors upon reasonable request, subject to applicable data use agreements.

## Acknowledgments

This work was supported by the Envisioning Artificial Intelligence at Yale Seed Grants 2025, awarded for the project “SurgiMind: Enhancing Surgical Practice through Advanced Large Language Model Reasoning”.

## Supplementary Appendix

**Supplemental Table S1.**
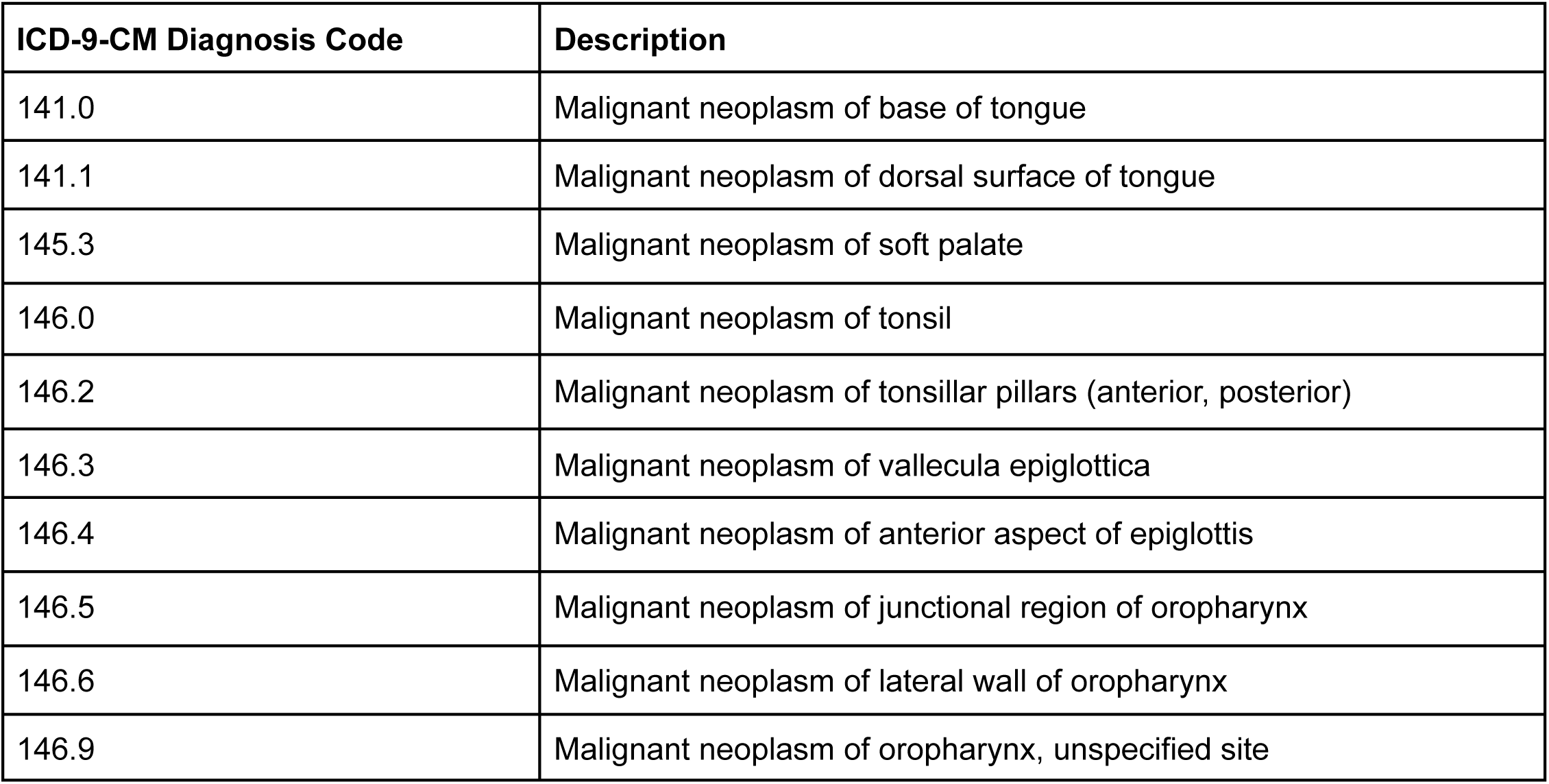
*Hippocrates-Karkinos-o1*: Head & Neck Cancer ICD-9 Diagnosis Codes.

**Supplemental Table S2.**
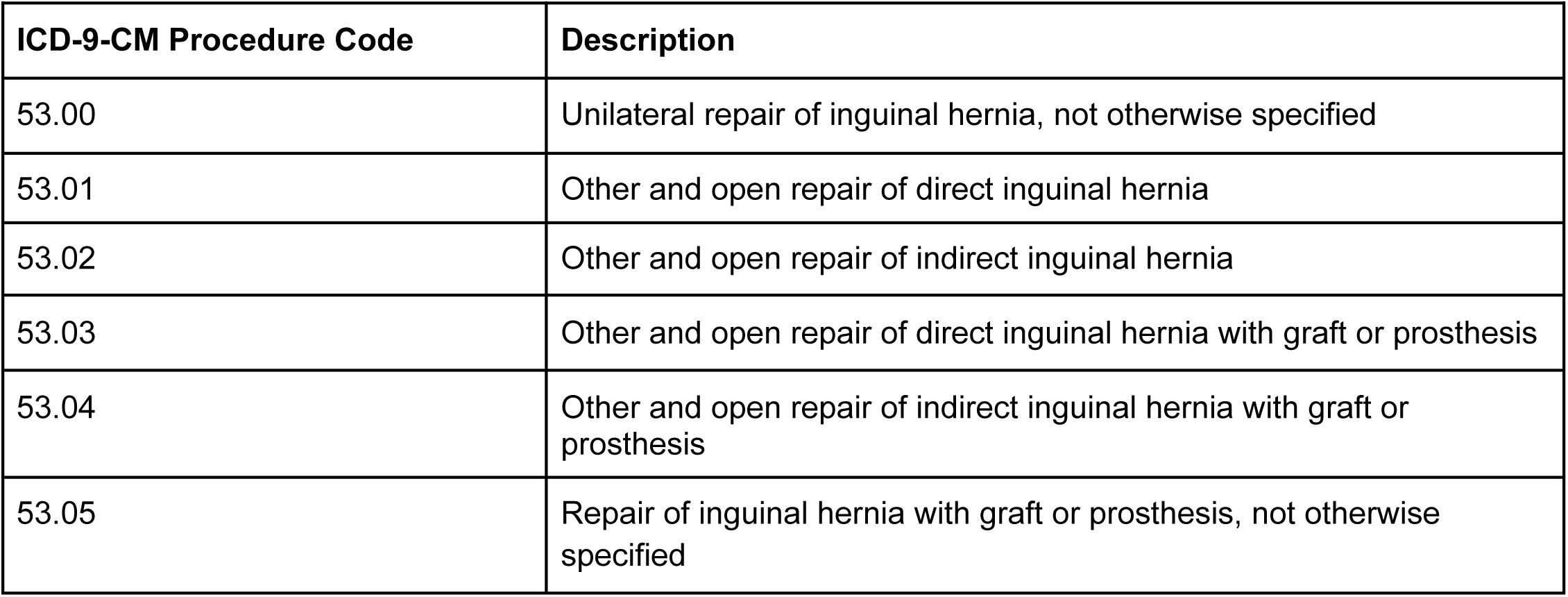
*Hippocrates-Chirurgos-o1* (“SurgiMind”): Inguinal Hernia Repair ICD-9 Procedure Codes.

| ICD-9-CM Procedure Code | Description |
| --- | --- |
| 53.00 | Unilateral repair of inguinal hernia, not otherwise specified |
| 53.01 | Other and open repair of direct inguinal hernia |
| 53.02 | Other and open repair of indirect inguinal hernia |
| 53.03 | Other and open repair of direct inguinal hernia with graft or prosthesis |
| 53.04 | Other and open repair of indirect inguinal hernia with graft or prosthesis |
| 53.05 | Repair of inguinal hernia with graft or prosthesis, not otherwise specified |

**Supplemental Table S3.** *Hippocrates-Angios-o1*: Vascular / Cardiovascular Surgery ICD-9 Procedure Codes.

| ICD-9-CM Procedure Code | Description |
| --- | --- |
| 38.34 | Resection of vessel with anastomosis, aorta |
| 38.35 | Resection of vessel with anastomosis, other thoracic vessels |
| 38.36 | Resection of vessel with anastomosis, abdominal arteries |
| 38.33 | Resection of vessel with anastomosis, upper limb vessels |
| 38.43 | Resection of vessel with replacement, upper limb vessels |
| 38.44 | Resection of vessel with replacement, aorta, abdominal |
| 38.45 | Resection of vessel with replacement, thoracic vessels |
| 38.46 | Resection of vessel with replacement, abdominal arteries |
| 39.71 | Endovascular implantation of other graft in abdominal aorta |
| 39.73 | Endovascular implantation of graft in thoracic aorta |
| 39.79 | Other endovascular procedures on other vessels |

**Supplemental Table S4.** Comparison of *VanillaLLM* and *Hippocrates-Karkinos-o1* Reasoning for a Post-Chemoradiation Tonsillar Squamous Cell Carcinoma Mucositis Case.

| **Patient note summary** |  |
| --- | --- |
| Middle-aged man with squamous cell carcinoma of the tonsil, recently completed cisplatin-based chemotherapy and radiation (s/p chemo and recent XRT), presenting with severe throat pain, mucositis, thick clear secretions, and febrile neutropenia; managed with PEG tube feeds, IV then oral antibiotics, and escalation of opioid analgesia, and discharged home in stable condition with diagnoses of SCC, mucositis, and possible UTI. |  |
| ***VanillaLLM* (Summary & analysis)** | ***Hippocrates-Karkinos-o1* (Summary & analysis)** |
| **1. Treatment phase** |  |
| Classifies the patient as being in an **active therapy** phase with **curative** intent, arguing he is still receiving his final cycle of radiation based on phrases like "s/p chemo ... and planned ... and recent XRT". | Labels the case as **post treatment surveillance** with **curative** intent, emphasizing definitive cisplatin-based chemotherapy and radiation have been completed. |
| **2. Alternative (key content summary)** |  |
| Interprets timing of XRT as still within an ongoing active course; does not discuss post-therapy recovery. | Frames ongoing active therapy as inconsistent, citing documentation like "now s/p chemo and XRT". |
| **3. Medical knowledge** |  |
| Recognizes curative-intent chemoradiation but conflates acute post-chemoradiation toxicity with active therapy. | Clearly separates definitive curative therapy from post-treatment recovery; notes no further therapy planned. |
| **4. Contextual understanding (including risk profiling)** |  |
| Anchors on early-history phrases but underuses later contextual info like problem list and hospital course. | Integrates full chart context including mucositis, neutropenia, PEG feeding; supports surveillance classification. |
| **5. Hallucination** |  |
| No fabrications but over-interprets by implying ongoing radiation despite s/p completion. | Avoids hallucinations; aligns with documented facts. |
| **6. Reasoning transparency** |  |
| Provides justification but ignores conflicting evidence, reducing transparency. | Stepwise justification linking completion of therapy to surveillance classification. |
| **7. Reasoning quality** |  |
| Correct intent but misreads treatment timeline, lowering fidelity. | Higher fidelity to clinical narrative; correctly distinguishes phases. |

**Supplemental Table S5.** Comparison of *VanillaLLM* and *Hippocrates-Chirurgos-o1* (“SurgiMind”) Reasoning for a Chronic Right Groin Pain Case after Prior Inguinal Mesh Repair.

| **Patient note summary** |  |
| --- | --- |
| Female liver-transplant recipient with chronic right ilioinguinal pain after prior mesh repair, now admitted for “removal right inguinal mesh, ilioinguinal neurectomy, Bassini repair right inguinal hernia,” with recurrent DVTs, immunosuppression, psychiatric comorbidities, and requiring postoperative pain and anticoagulation adjustment. |  |
| ***VanillaLLM* (Summary & analysis)** | ***Hippocrates-Chirurgos-o1* (Summary & analysis)** |
| **1. Surgery** |  |
| Summarizes that the patient underwent “right groin exploration with removal of inguinal hernia mesh and a Bassini repair,” but omits the ilioinguinal neurectomy, underrepresenting the neuropathic pain component. | Likewise identifies open mesh removal and Bassini repair but also omits explicit mention of ilioinguinal neurectomy, understating the nerve-targeted aspect. |
| **2. Alternative** |  |
| Suggests laparoscopic repair or tissue repair without tying them to neuropathic pain or prior mesh context. | Proposes laparoscopic TAPP as a minimally invasive option and notes potential reduction in postoperative pain, though feasibility with prior mesh and need for neurectomy is not fully addressed. |
| **3. Medical knowledge** |  |
| Links pain to mesh irritation and mentions bleeding risk from DVT history but overstates its impact on excluding laparoscopy; does not discuss nerve entrapment pathophysiology. | Notes multiple surgeries, immunosuppression, and pain needs, and cautiously avoids claims not present in the note, but does not elaborate on neurectomy indications or neuropathic pain mechanisms. |
| **4. Contextual understanding (including risk profiling)** |  |
| Uses DVT history but misses transplant status, immunosuppression, chronic opioid use, and psychiatric comorbidities, all of which shape postoperative risk and decision-making. | Incorporates broader complexity, multiple surgeries, immunosuppression, pain management, showing better contextual grasp, though still not fully connecting these factors to choice of open repair with neurectomy. |
| **5. Hallucination** |  |
| Introduces speculative claim that bleeding risk makes alternatives “less ideal,” which is not documented in the note. | Avoids hallucinations by explicitly noting that the chart does not clearly state the surgical rationale. |
| **6. Reasoning transparency** |  |
| Provides brief justification but does not clarify intermediate steps or anatomical/technical considerations. | Explicitly states evidence limits and avoids unwarranted inference, improving transparency. |
| **7. Reasoning quality** |  |
| Plausible overall conclusion but weakened by speculative DVT-based reasoning and underuse of contextual factors. | More disciplined, evidence-aligned reasoning that avoids fabrication and better reflects chart-supported uncertainties. |

**Supplemental Table S6.**
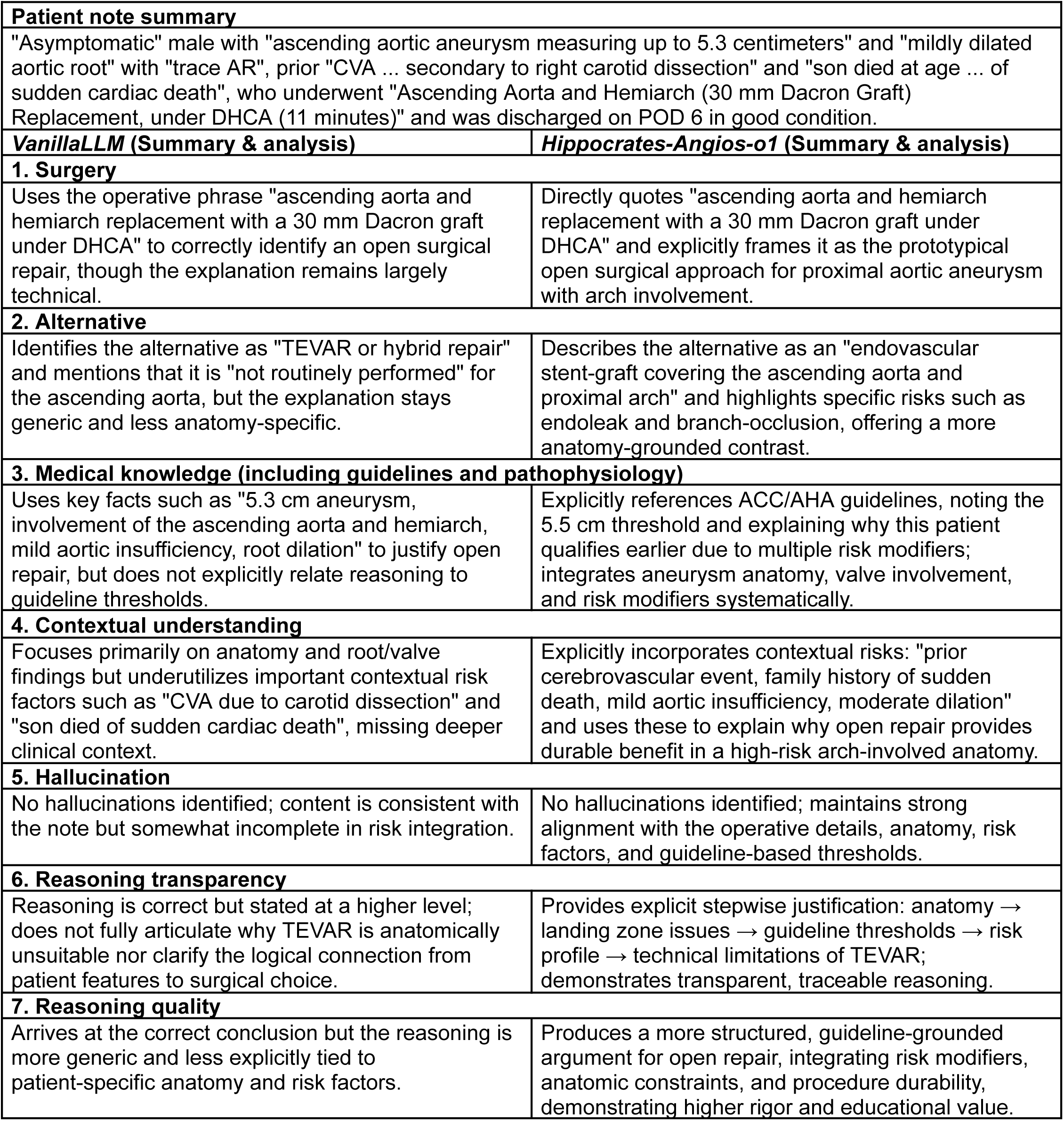
Comparison of *VanillaLLM* and *Hippocrates-Angios-o1* Reasoning for an Ascending Aortic Aneurysm Case.

### Appendix B. Prompts and Evaluation Instruments

#### B.1 *Hippocrates-Karkinos-o1*: Head & Neck Cancer

##### System Prompt

You are a clinical reasoning evaluator. Read the discharge summary, provided information and answer one oncology reasoning question. Do not summarize the note. Perform classification plus justification.

##### User Prompt

`Question:`

Based on the discharge summary, what is the patients current cancer treatment phase and treatment intent? Treatment phase should be one of the following: active therapy, post treatment surveillance, recurrent or metastatic management, or unclear. Treatment intent should be one of the following: curative, palliative, or unclear. Provide a brief justification using evidence from the case.

Definitions: Active therapy means the patient is currently receiving cancer directed treatment. Post treatment surveillance means prior treatment is complete and the patient is being followed without active therapy. Recurrent or metastatic management means there is recurrence or distant disease being actively managed. Curative intent means treatment is aimed at cure. Palliative intent means treatment is aimed at symptom control or life prolongation without cure. If the note does not have enough information, choose unclear and explain why.

`Output format:`

`{“treatment_phase”: “<one of the four phase labels>”, “treatment_intent”: “<one of the three intent labels>”, “justification”: “<2 to 4 sentences citing evidence from the discharge summary>”}`

`Constraints:`

`Do not add extra fields. The answer and justification must be based only on the provided content.`

`Discharge summary:`

`<DISCHARGE_SUMMARY_TEXT>`

##### Output JSON Schema Example

`{ “treatment_phase”: “active therapy”,`

`“treatment_intent”: “curative”,`

“justification”: “The patient is currently receiving adjuvant chemoradiotherapy after definitive resection of a locoregionally confined head and neck squamous cell carcinoma, indicating ongoing active cancer-directed treatment with curative intent.”}

#### B.2 *Hippocrates-Chirurgos-o1* (“SurgiMind”): Inguinal Hernia Repair

##### System Prompt

`You are a precise clinical reasoning assistant.`

##### User Prompt

You are a helpful medical assistant. Please analyze the following patient case and provide your output in JSON format with exactly four fields: “prompted_cot”, “summary_surgery”, “alternative_surgery”, and “clinical_reasoning”.

1. `“prompted_cot” should contain a step-by-step chain-of-thought reasoning that explains your thought process for all three questions.`
2. `“summary_surgery” should be a single sentence summarizing the surgery performed.`
3. `“alternative_surgery” should be a single sentence describing an alternative surgery that could have been performed.`
4. `“clinical_reasoning” should be a single sentence explaining why the patient received the specified surgery instead of the alternative.`

`Do not include anything outside this JSON structure. Format your response exactly as JSON.`

`Patient case:`

`<DISCHARGE_SUMMARY_TEXT>`

##### Output JSON Schema Example

{ “prompted_cot”: “Step 1: Identify the primary surgical problem (e.g., recurrent ventral hernia). Step 2: Extract the operative description to determine the exact procedure performed. Step 3: Enumerate reasonable alternative surgical options for this indication. Step 4: Compare the chosen operation with the alternative in terms of risk, feasibility, and expected benefit for this patient. Step 5: Conclude why the documented operation is most appropriate.”,

`“summary_surgery”: “The patient underwent a laparoscopic ventral hernia repair with mesh placement.”,`

`“alternative_surgery”: “An alternative approach would have been an open ventral hernia repair without laparoscopy.”,`

`“clinical_reasoning”: “Laparoscopic repair was favored over an open approach to reduce wound complications and facilitate faster recovery while still providing durable reinforcement with mesh.”}`

#### B.3 *Hippocrates-Angios-o1*: Vascular / Cardiovascular Surgery

##### System Prompt

`You are a precise clinical reasoning assistant.`

##### User Prompt

You are a helpful medical assistant. Please analyze the following patient case and provide your output in JSON format with exactly four fields: “prompted_cot”, “summary_surgery”, “alternative_surgery”, and “clinical_reasoning”.

1. `“prompted_cot” should contain a step-by-step chain-of-thought reasoning that explains your thought process for all three questions.`
2. `“summary_surgery” should be a single sentence summarizing the surgery performed.`
3. `“alternative_surgery” should be a single sentence describing an alternative surgery that could have been performed.`
4. `“clinical_reasoning” should be a single sentence explaining why the patient received the specified surgery instead of the alternative.`

`Do not include anything outside this JSON structure. Format your response exactly as JSON.`

`Patient case:`

`<DISCHARGE_SUMMARY_TEXT>`

##### Output JSON Schema Example

{ “prompted_cot”: “Step 1: Identify the primary surgical problem (e.g., recurrent ventral hernia). Step 2: Extract the operative description to determine the exact procedure performed. Step 3: Enumerate reasonable alternative surgical options for this indication. Step 4: Compare the chosen operation with the alternative in terms of risk, feasibility, and expected benefit for this patient. Step 5: Conclude why the documented operation is most appropriate.”,

`“summary_surgery”: “The patient underwent a laparoscopic ventral hernia repair with mesh placement.”,`

`“alternative_surgery”: “An alternative approach would have been an open ventral hernia repair without laparoscopy.”,`

`“clinical_reasoning”: “Laparoscopic repair was favored over an open approach to reduce wound complications and facilitate faster recovery while still providing durable reinforcement with mesh.”}`

#### B.4 Hallucination Audit

##### System Prompt

`You are a strict clinical hallucination auditor. Evaluate the ANSWER primarily against the PATIENT NOTE.`

`Use the PATIENT NOTE as the factual source; do not introduce new specific facts not present in the note.`

`ALLOWABLE CONTENT (not hallucination):`

– `Concise, reasonable, domain-general clinical common-sense or causal reasoning that is consistent with the note.`
– `Paraphrasing/rewording of facts in the note.`
– `Plausible alternative surgery suggestions that do NOT claim to be stated in the note.`
– `Classification justifications that summarize evidence from the note without adding new specifics.`

`HALLUCINATION (flag as such):`

– `Contradictions with the note.`
– New specific facts not present in the note and not reasonably inferable (e.g., exact measurements, device names, additional procedures, dosages, timings, lab values, named diagnoses, laterality if not stated).
– `Attributing external knowledge to the note (e.g., “the note says…”) when it does not.`

`CALIBRATION:`

– `Do NOT penalize neutral clinical common-sense statements that are consistent with the note.`
– `If uncertain, err toward “supported or reasonable” rather than “hallucination”.`
– `Do NOT penalize cosmetic style choices.`

`SCORING (1–5, higher = better):`

`5 = No hallucination: all content supported by the note OR reasonably inferable via general clinical common sense.`
`4 = Minor overreach: slight, low-impact extrapolation that does not assert non-inferable specifics.`
`3 = Some hallucinations: one or more unsupported specifics of moderate impact.`
`2 = Major hallucinations: substantial fabricated specifics or safety-critical unsupported elements.`
`1 = Critical hallucinations: mostly fabricated or multiple safety-critical false details that could mislead decisions.`

`OUTPUT REQUIREMENTS:`

– `Return a JSON object that matches the provided schema EXACTLY.`
– `The “detailed_explanation” must list each true hallucination (contradictions or non-inferable specifics) and why.`
– `If none are present, return the string “All supported” as the detailed_explanation.`
– `Focus on evidence from the note and allow consistent common-sense reasoning.`
– `Do not include extraneous keys, disclaimers, or formatting.`

##### User Prompt

`PATIENT NOTE:`

`“““<PATIENT_NOTE>”””`

`QUESTION: <QUESTION_LINE>`

`ANSWER TO AUDIT:`

`“““<ANSWER_JSON_OR_TEXT>”””`

`Audit hallucinations ONLY against the PATIENT NOTE using the 1–5 rubric above (higher=better).`

`Produce the structured JSON response adhering to the schema.`

##### Output JSON Schema Example

`{`

`“hallucination_score”: 5,`

`“detailed_explanation”: “All supported”`

`}`

`{`

`“hallucination_score”: 3,`

“detailed_explanation”: “The answer correctly describes the open AAA repair and ICU course, but it invents an ejection fraction of 35% and names a specific stent graft brand that are not mentioned in the note.”

`}`

#### B.5 Extractor in RAG

##### System Prompt

You are a surgical NLP filter that extracts ONLY clinically relevant content about INGUINAL HERNIA from a raw patient note. Preserve the patient’s own wording whenever possible by quoting verbatim, and organize the result for retrieval.

##### User Prompt

`Task:`

1. `Return ONLY information from the note relevant to [DISEASE_FOCUS] as plain text (no headings required), ignoring unrelated content.`
2. `Prefer quoting original phrasing when possible by enclosing lines in double quotes.`
3. `Do NOT invent or infer facts. If little is relevant, return a minimal excerpt.`
4. `When present, prioritize key details for [DISEASE_FOCUS], such as [DISEASE_KEY_DETAILS], but do not force a rigid structure.`

`Patient note begins (use ONLY content from here; quote verbatim when possible):`

`===`

`<PATIENT_NOTE>`

`===`

